# Serological Response in Lung Transplant Recipients after Two Doses of SARS-CoV-2 mRNA Vaccines

**DOI:** 10.1101/2021.04.26.21255926

**Authors:** Madhusudhanan Narasimhan, Lenin Mahimainathan, Andrew E. Clark, Amena Usmani, Jing Cao, Ellen Araj, Fernando Torres, Ravi Sarode, Vaidehi Kaza, Chantale Lacelle, Alagarraju Muthukumar

## Abstract

The immunological effectiveness of SARS-CoV-2 mRNA vaccines in lung transplant (LT) recipients is largely unknown. Thus, we assessed the effect of Pfizer-BioNTech and Moderna mRNA vaccines’ two-dose (2D) regimen on humoral responses in immunocompromised lung transplant (LT) recipients. About 25% (18/73) of SARS-CoV-2 uninfected-LT patients generated positive spike-IgG response following 2D of vaccines, with 36% (9/25) in the Moderna cohort and only 19% (9/48) in the Pfizer cohort. 2D in LT patients elicited significantly lesser median IgG_SP_ response (1.7 AU/mL, 95% CI: 0.6-7.5 AU/mL) compared to non-transplanted, uninfected naïve subjects (14209 AU/mL, 95% CI: 11261-18836 AU/mL) (p<0.0001). In LT patients, Moderna-evoked seropositivity trend was higher by 23-fold than Pfizer. 2D COVID-19 vaccination elicits a dampened serological response in LT patients. Whether assessing other arms of host immunity combined with higher vaccine dose can better capture and elicit improved immunogenicity in this immunocompromised population warrants investigation.

---

Lung-transplant (LT) recipients are at high risk for severe COVID-19 due to immunosuppression (IS) and respiratory tropism for SARS-CoV-2. Although single dose SARS-CoV-2 vaccine-related immunogenicity in solid organ transplant have emerged^1^, so far there has been no evidence of the impact of 2-dose vaccine regimen on the quality of humoral immunological response in LT recipients. Understanding the impact of vaccine regimens on humoral immunity is critical to optimize the COVID-19 immunization in this immunocompromised cohort.

## Methods

Serum samples from LT recipients vaccinated for SARS-CoV-2 with 2D of either the Pfizer-BioNTech or Moderna vaccines or 2D-vaccinated naïve (non-transplanted and COVID-19 non-exposed) group were used in this study. They were de-identified, discarded, and remnant blood samples available in the laboratory after routine analysis. This study involved no specific collection of samples. The University of Texas Southwestern Medical Center’s Institutional Review Board granted a waiver of consent for this study.

Antibody responses were semi-quantitatively assessed using serum samples analyzed on the Alinity i platform (Abbott Laboratories, Abbott Park, IL) using the FDA-approved SARS-CoV-2 anti-nucleocapsid protein IgG assay (IgG_NC_), the SARS-CoV-2 anti-spike protein IgM assay (IgM_SP_), or the SARS-CoV-2 anti-spike protein IgG II assay (IgG_SP_) as previously described.^2^ Index values of ≥1.4 (IgG_NC_), ≥ 1.0 (IgM_SP_), and ≥ 50 AU/ml (IgG_SP_) were interpreted as positive per the manufacturer’s recommended threshold. IgG_NC_ positivity informs natural SARS-CoV-2 infection, while IgG_SP_/IgM_SP_ positivity strongly correlate with emergence of natural or vaccine-driven neutralizing immunity.^2,3^ CD4+ T-cell activity was assessed as a marker of immune competence using the ImmuKnow® assay (Viracor-IBT). Results were interpreted as either low, moderate, or strong correlating with manufacturer established ATP ranges of ≤ 225, 226-524, and ≥ 525 ng/mL, respectively. Two-tailed unpaired t-test was performed to assess statistical significance using GraphPad Prism 9.1.0. A value of p<0.05 was considered statistically significant.

## Results

There were 73 LT recipients included and samples were collected and analyzed between February 1, 2021 and March 19, 2021 (Table 1). The median age was 65 years (Interquartile range [IQR], (53.5-69.5 years)), 74% were male. 66% received Pfizer vaccine, 34% received the Moderna formulation. LT were on standard IS with tacrolimus, anti-metabolite and steroids as per institutional protocol. On review, one recipient was not on anti-metabolite, two were on cyclosporine and one was on combination low dose tacrolimus and sirolimus. Median time since transplant surgery (TSTS) for study participants was 40 months [IQR, 44 (19-63 months)]. IgG_NC_ assessment confirmed the absence of any previous silent or asymptomatic COVID-19 infection in naïve, non-transplant cohorts with one previously infected case in the LT cohort.

**Table 1:**
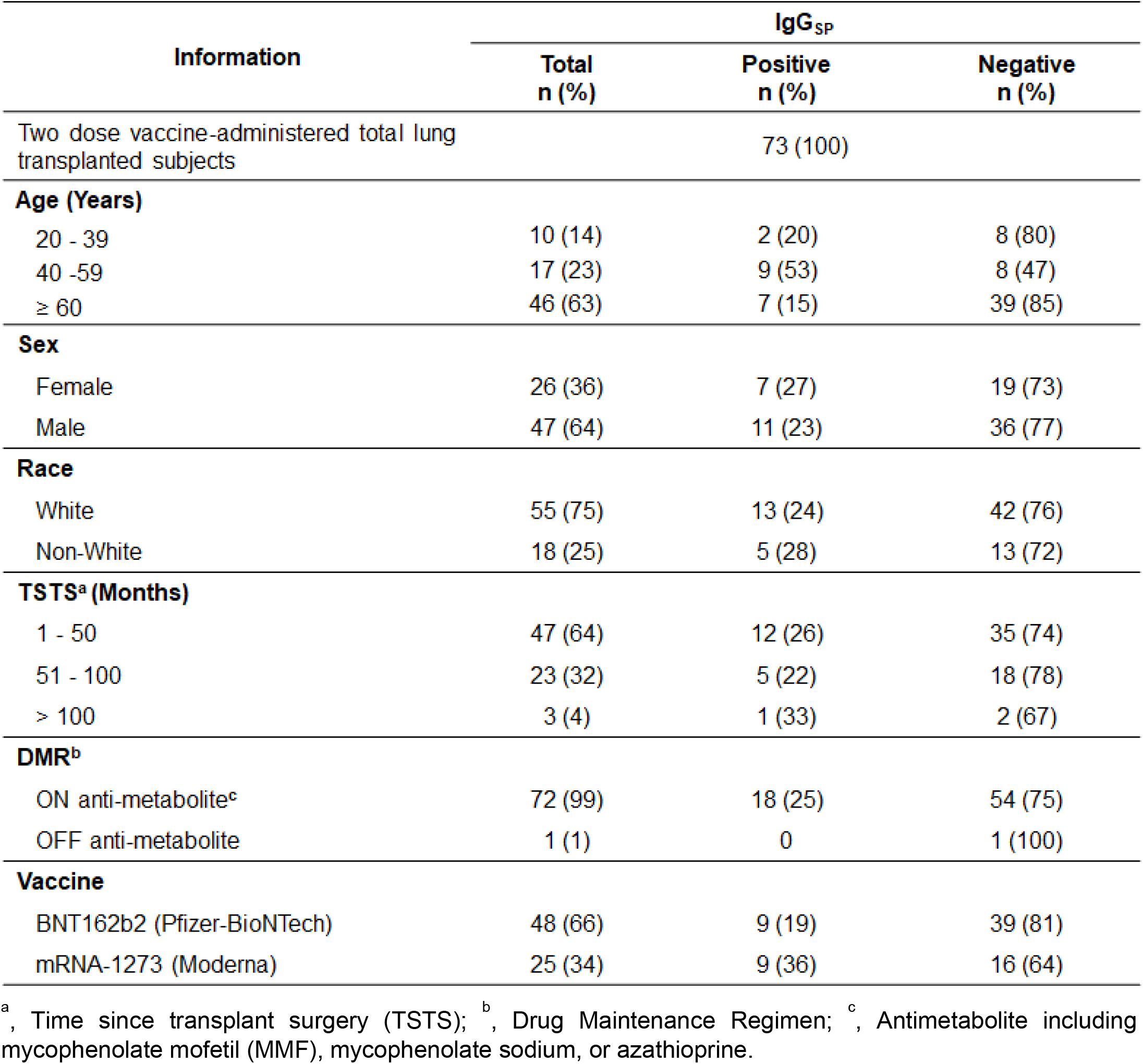
Demographic and Clinical Characteristics of Study Participants, Stratified by Spike-specific IgG (IgG_SP_) Antibody Response after Two Dose-COVID-19 Vaccination.

Only 25% (18/73) of LT recipients demonstrated IgG_SP_-positivity following 2D of mRNA vaccines. At a median of 17.5 days after the second dose of the Pfizer vaccine, only 19% (9/48) displayed positive IgG_SP_ levels. In contrast, at a median time of 19 days after 2D of Moderna-mRNA vaccine, 36% (9/25) had IgG_SP_-positivity (Table 1; Fig. 1).

**Figure 1:**
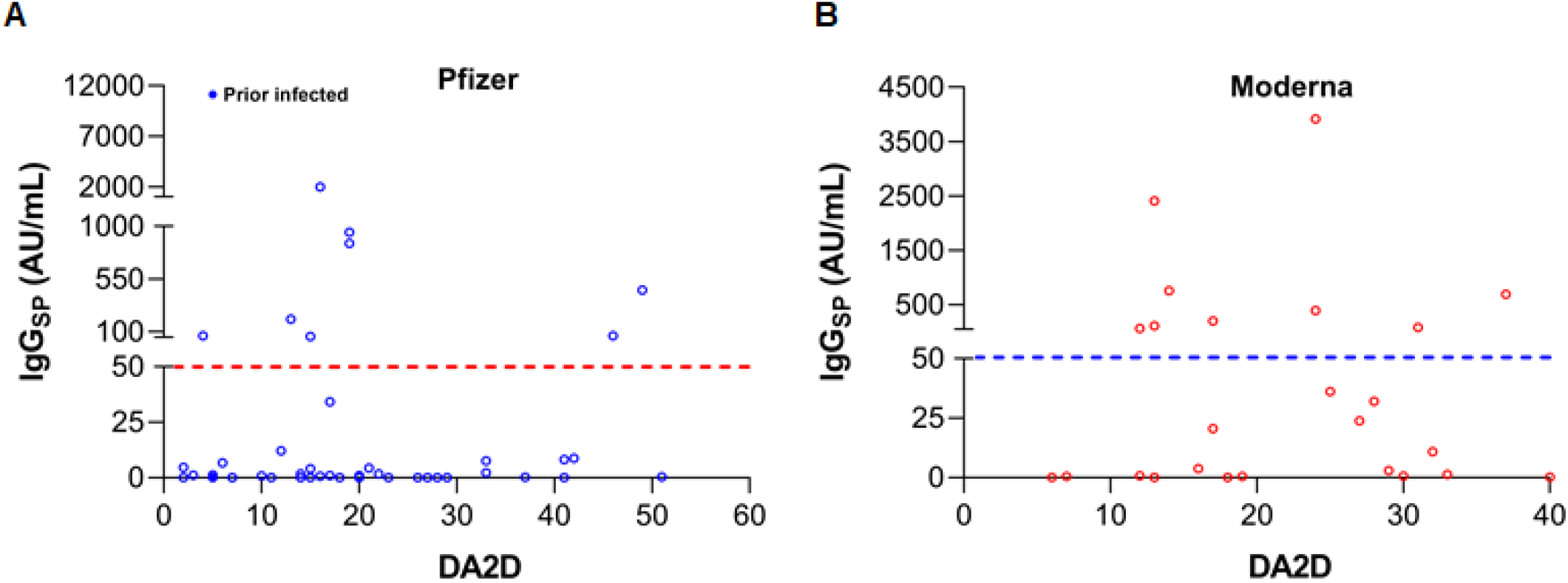
Frequency Distribution of IgG_SP_ versus Days after Second Dose of COVID-19 Vaccines in Lung Transplant Recipients (A) BNT162b2 (Pfizer-BioNTech); (B) mRNA-1273 (Moderna). DA2D, Days after Second Dose of COVID-19 Vaccines; Dotted red and blue lines, IgG_SP_ serology assay’s manufacturer recommended positive cut-off value.

56 among 73 LT recipients had Cylex Immuknow assay values measured. Moderna vaccine was found to elicit positive IgG_SP_ responses by 44% (4/9) and 50% (1/2) in the moderate and strong category, respectively. In contrast, the Pfizer vaccine elicited a positive IgG_SP_ response only in 18% (3/17) and none (0/6) in patients with moderate and strong ImmuKnow levels, respectively (Table S1). There was no trend for better antibody response for IgG_SP_, IgM_SP_, or IgG_NC_ based on Cylex Immuknow assay in both groups.

Comparison of SARS-CoV-2 specific antibody responses following a 2D regimen of mRNA vaccine in LT recipients to non-transplant, immunologically naïve (never SARS-CoV-2 infected) participants clearly illustrated that the median IgG_SP_ levels were significantly lower among immunosuppressed-LT recipients (median of 1.7 AU/mL, 95% CI: 0.6-7.5 AU/mL) compared to naïve individuals (median of 14209 AU/mL, 95% CI: 11261-18836 AU/mL) (p<0.0001; Fig. 2). Furthermore, albeit statistically non-significant, a lower circulating IgG_SP_ trend with 2D-Pfizer vaccine was noted than the 2D-Moderna formulation among LT patients (Fig. S1; 23-fold, p=0.9555).

**Figure 2:**
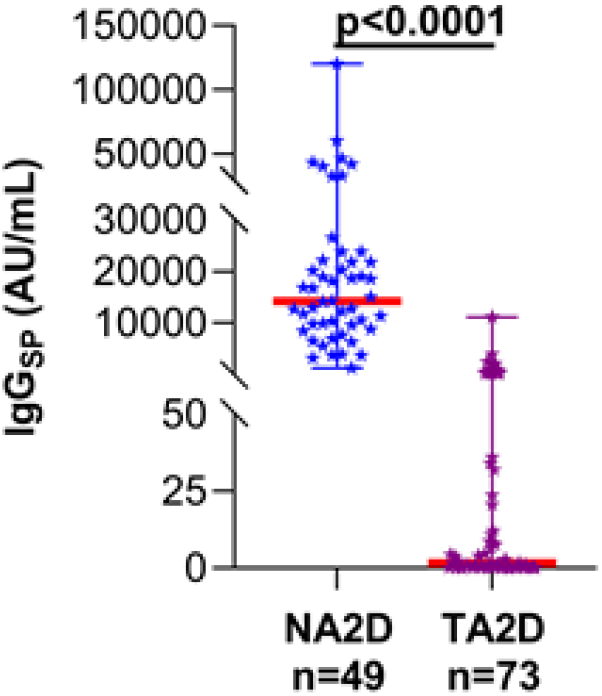
IgG_SP_ levels in Naïve Subjects and Lung Transplant Patients after 2 doses of COVID-19 vaccine. NA2D, Naïve Subjects (non-transplanted and not had previous SARS-CoV-2 infection) after 2 doses of Vaccine; TA2D, Lung Transplant Patients after 2 doses of Vaccine; Thick red line, median value.

## Discussion

This is the first study to evaluate the immunogenicity of the Pfizer and Moderna COVID-19 vaccines following the 2D regimen in LT recipients. Our data shows that only a minority (25%) of the participants mounted appreciable anti-spike antibody responses. In addition, the ImmuKnow Cylex assay-based results suggested that the immune cell function-based stratification does not predict antibody response to the vaccines. However, irrespective of the immune cell function, the Moderna formulation generated a trend towards more robust antibody response than the Pfizer vaccine in immunosuppressed-LT patients, indicating an enhanced protective immunity. The principle of generating a mRNA vaccine that encodes a SARS-CoV-2 spike protein that is stabilized in the prefusion conformation along with the larger dose of 100 µg in the Moderna versus the 30 µg in the Pfizer preparation could explain the differential antibody response obtain when comparing the two vaccines.^4,5^ Similar findings were reported recently^1^ with Moderna group at developing antibody response at 69% vs 31% in Pfizer group.

Importantly, our data indicates that receiving two doses of vaccine does not mean an assured protection against COVID-19 infection for the majority of LT patients, as majority exhibit either nil or negligible IgG_SP_ responses. Evidence from a non-COVID19 context supports the doubling of vaccine dose as an attractive strategy to improve the immunogenicity in transplant recipients.^6^ But the theoretical risk of vaccines and alloimmunity and rejection remains, although not a consistent observation so far. It also remains to be evaluated if such lower antibody response early after vaccination would suffice to mitigate risk for COVID-19.

Limitations include small sample size, lack of demographic data in non-transplant group, absence of serial measurements after vaccination, and shorter time for follow up. Despite these limitations, we demonstrate that a two-dose format of COVID-19 mRNA vaccines elicit a serological response, though not in a majority of LT patients. Further studies are needed to understand the efficacy and longevity of vaccine-derived COVID-19 immunity in this vulnerable population. Since most lung transplant recipients are maintained at higher level of immunosuppression compared to other solid organ transplant recipients, larger studies with longer duration of follow-up are needed to confirm our preliminary findings of antibody response after completing SARS-CoV-2 mRNA vaccination.

## Data Availability

Due to ethical reasons, the data are not publicly available. However, upon request, the data presented in this study can be shared.

## Author Contributions

Concept and Design: MN, LM, VK, CL, AM

Acquisition, analysis, or interpretation of data: MN, LM, EA, VK, AU, CL, AM

Statistical analysis and verification: MN, LM, VK, AM

Drafting of the manuscript: MN, LM, AM

Critical revision of the manuscript for important intellectual content: MN, LM, AEC, RS, VK, CL, FT, AM

Administrative, technical, or material support: RS, CL, AM

Supervision: CL, AM

## Additional Contributions and Acknowledgments

We thank our medical technologists, Charles Alexis and Kimberly Fankhauser for helping with laboratory testing. We also thank Ashley Comeaux, Ryan Osterberg, Katrina Bolls, and Anthony Spinelli for retrieving samples, labeling tubes, and aliquoting. We acknowledge Marcie Buford for pulling up the list of vaccinated patients. We thank Abbott’s diagnostic division for providing reagents.

## Figure Legends

Figure S1: **Evaluation of Immunogenicity following double dose Pfizer and Moderna Vaccine Regimen in LT Recipients**. Thick red line, median value; S4A, Median (95% CI) [Pfizer – 0.9 (0.0-4.1); Moderna -20.6 (0.8-80.2)].

Table S1: **IgM**_**SP**_, **IgG**_**NC**_, **and IgG**_**SP**_ **Responses following Double Dose COVID-19 Vaccine Regimen in Lung Transplant Patients, Stratified by Cylex ImmuKnow assay levels**. + and -, Serology assay’s positive and negative results based on the manufacturer recommended corresponding cut-off value.

