## Supplemental Table 1 and Fig. s1 for "Serological Response in Lung Transplant Recipients after Two Doses of SARS-CoV-2 mRNA Vaccines"

Madhusudhanan Narasimhan, PhD, MSHA[1#], Lenin Mahimainathan, PhD, MB(ASCP)[1#], Andrew E. Clark, PhD, DABMM[1], Amena Usmani, MD[1], Jing Cao, PhD, DABCC[1], Ellen Araj, MD[1], Fernando Torres, MD[2], Ravi Sarode, MD[1,2], Vaidehi Kaza, MD, MPH[2], Chantale Lacelle, PhD, D(ABHI)[1*], Alagarraju Muthukumar, PhD, DABCC[1*]

[1] Department of Pathology, University of Texas Southwestern Medical Center, Dallas, Texas

[2] Department of Internal Medicine, University of Texas Southwestern Medical Center, Dallas, Texas

[#] - Equal Contribution

***Corresponding author:**

Alagarraju Muthukumar, PhD, DABCC or Chantale Lacelle, PhD, DABHI

UT Southwestern Medical Center, Dallas

 (AM) (or)

 (CL)

Ph: 214-645-5103 (AM)/214-648-0904 (CL)

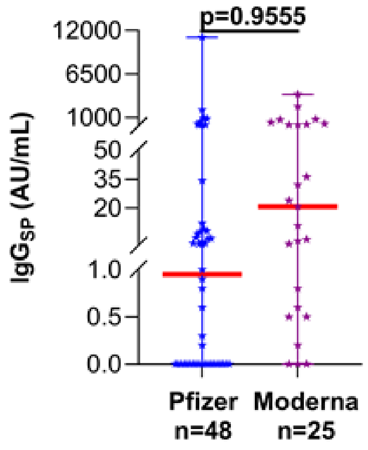
p=0.9555
12000
6500
1000
50
35
20
1.0
0.5
0.0
IgG$_{SP}$ (AU/mL)
Pfizer
n=48
Moderna
n=25

**Figure S1**

**Table S1: $IgM_{SP}$, $IgG_{NC}$, and $IgG_{SP}$ Responses following Double Dose COVID-19 Vaccine Regimen in Lung Transplant Patients, Stratified by Cylex ImmuKnow assay levels**

| Information | n (%) | Cylex ImmuKnow assay levels | | |
|---|---|---|---|---|
| | | Low n (%) | Moderate n (%) | Strong n (%) |
| Two dose vaccine administered lung transplanted total subjects that had Cylex results recorded | 56/73 (77) | 22/56 (39.3) | 26/56 (46.4) | 8/56 (14.3) |
| | **$IgG_{SP}$ serology results** | | | |
| BNT162b2 (Pfizer-BioNTech) | + | 3/15 (20) | 3/17 (18) | 0/6 (--) |
| | – | 12/15 (80) | 14/17 (82) | 6/6 (100) |
| mRNA-1273 (Moderna) | + | 1/7 (14) | 4/9 (44) | 1/2 (50) |
| | – | 6/7 (86) | 5/9 (56) | 1/2 (50) |
| | **$IgM_{SP}$ serology results** | | | |
| BNT162b2 (Pfizer-BioNTech) | + | 1/15 (7) | 1/17 (6) | 0/6 (--) |
| | – | 14/15 (93) | 16/17 (94) | 6/6 (100) |
| mRNA-1273 (Moderna) | + | 0/7 (--) | 0/9 (--) | 0/2 (--) |
| | – | 7/7 (100) | 9/9 (100) | 2/2 (100) |
| | **$IgG_{NC}$ serology results** | | | |
| BNT162b2 (Pfizer-BioNTech) | + | 0/15 (--) | 1/17 (6) | 0/6 (--) |
| | – | 15/15 (100) | 16/17 (94) | 6/6 (100) |
| mRNA-1273 (Moderna) | + | 0/7 (--) | 0/9 (--) | 0/2 (--) |
| | – | 7/7 (100) | 9/9 (100) | 2/2 (100) |

+ and — , Serology assay's positive and negative results based on the manufacturer recommended corresponding cut-off value.
